# The use of heart rate monitoring in patient-caregiver interactions: a protocol for a mixed-methods study on feeling of closeness and safety

**DOI:** 10.1101/2025.11.03.25339169

**Authors:** Maria Madsen, Bjørnar Hassel, Daniel S. Quintana, Emilie S. M. Kildal

## Abstract

**Background:** Patients with profound intellectual and multiple disabilities (PIMD) are unable to communicate verbally, which makes it challenging for caregivers to understand and detect their emotions and needs. HR monitoring has been used as a tool to potentially gain a better understanding, as it can help identify distress and pain in these patients. Despite this, little is known about how HR monitoring affects relational aspects of care, including closeness and safety from the caregiver’s perspective.

**Objectives:** This study aims to investigate how the use of HR monitoring as a communicative tool in everyday care affects caregivers’ perceived closeness in their interactions with patients with PIMD.

**Methods:** *Participants*: Caregivers who work regularly with patients with PIMD and who participate in the clinical testing of HR monitoring in care.

**Design:** The study uses a mixed-methods design. Pre-post questionnaires from approximately 58 caregivers using The Non-Verbal Patient-Caregiver Understanding Questionnaire and study-specific items will be used for quantitative data on closeness and safety. Semi-structured interviews with a subset of 4-6 caregivers will be carried out to collect qualitative data. Qualitative data will be analyzed using reflexive thematic analysis, while quantitative data will be analyzed using within-subject paired samples t-tests and regression analyses.

**Discussion:** The study will advance our understanding of how HR monitoring affects relational aspects of care, in particular closeness and safety.

## Introduction

Intellectual disability (ID) is a neurodevelopmental condition characterized by significant limitations in intellectual functioning and adaptive behavior, with onset before the age of 18 (American Psychiatric Association, 2013; World Health Organization, 2019/2021). ID affects approximately 1% of the population (Zablotsky et al., 2019; Maulik et al., 2011). Severity ranges from mild to profound, with about 5% of individuals falling within the severe (IQ 20– 34) or profound (IQ <20) range (van Bakel et al., 2014; Smith et al., 2020). These individuals typically require lifelong, round-the-clock care. ID can arise from acquired causes such as hypoxia, ischemia, or infection early in life (Conway et al., 2018; Ricci et al., 2008; de Montferrand et al., 2019; Ostrander & Bale, 2019), but genetic syndromes and de novo mutations are frequent causes (Martin et al., 2018).

Profound ID is often accompanied by additional neurological and somatic conditions. Cerebral palsy (CP), which affects posture, movement, and oromotor control, is common in this population (Reid et al., 2018). Epilepsy and other chronic somatic conditions are also prevalent (Zandbelt et al., 2024). Many individuals have combined impairments that place them in the category of profound intellectual and multiple disabilities (PIMD) (Mol-Bakker et al., 2024). PIMD is characterized by the coexistence of profound intellectual impairment and severe motor disabilities, often accompanied by sensory impairments and complex health needs (Matérne & Holmefur, 2022; van Timmeren et al., 2017).

People with PIMD struggle to communicate unequivocally. Their communication can consist of sounds, movements, facial expressions, muscle tension, and body posture rather than verbal language (Maes et al., 2007), making it difficult for caregivers to detect needs, distress, and pain (Regnard et al., 2007).

In general healthcare, effective communication is central to forming bonds and relations with patients, and it can benefit patient-centered outcomes (Sharkiya, 2023). Effective communication includes language and speech, in addition to non-verbal communication such as eye contact, touch, empathy, signals, listening, patience, time, and distance (Lacerda et al., 2021). Lacerda and colleagues (2021) also highlight that effective communication facilitates the formation of patient-caregiver relationships, interactions, and trust. People with PIMD typically need long-term care as a result of their complex needs and high dependency (van Cooten et al., 2022). For patients in long-term care, the caregiver’s relational behavior plays an important role in the patient’s well-being, behavior, and mood (McGilton et al., 2012). For patients with PIMD, emotional well-being, including feelings of closeness, safety, warmth, joy, and appreciation, is important for the quality of the interactions between caregivers and patients, and for the patient’s quality of life (Hostyn et al., 2011; Petry & Maes, 2007). Feelings of safety are also central for patients to optimally develop their abilities (Petry & Maes, 2007). Given this, the importance of the caregivers′ ability to interpret and understand the patient becomes more evident in relationships with patients with communication difficulties. This highlights the need for strategies and tools that can support caregivers′ ability to emotionally attune to patients with communication difficulties.

The general principles of communication are especially relevant in the context of PIMD, where caregivers face additional challenges. Professional caregivers play a central role in the daily lives of people with PIMD. Their responsibilities extend beyond physical care to include interpreting subtle expressions, preventing suffering, and fostering relational well-being (Hoogsteyns et al., 2023). The sense of closeness and emotional connection caregivers feel in their interactions is not only vital for patient quality of life (Nieuwenhuijse et al., 2020), but also for caregiver satisfaction, resilience, and prevention of compassion fatigue (Singh et al., 2020; Davenport & Zolnikov, 2022). Establishing closeness is particularly challenging when communication is minimal, and caregivers often report uncertainty about whether patients feel understood, safe, and emotionally connected (Kanyane & Maseko, 2024; Nieuwenhuijse et al., 2020; Penninga et al., 2022, Boysen et al., 2024).

In this study, we define *closeness* as the caregiver’s experience of emotional attachment and connection in interactions with the patient. It involves a sense of being attuned to the patient′s expressions and needs, feeling that they share a bond that is meaningful in the everyday interactions with the patient, experiencing understanding and warmth in the relationship.

In this study, *safety* refers to the caregiver′s sense of confidence in their own actions and judgements towards the patient. It does not entail physical safety, but rather the trust that one′s decisions and actions are what′s best for the patient. Safety describes the caregiver′s feeling of being secure and steady in their caregiving role.

Technological tools that detect physiological signs, such as heart rate (HR), may support caregivers in understanding minimally communicative patients. HR is defined by the frequency of heartbeats in a given time frame, most commonly reported by beats per minute (bpm) (Kranjec et al., 2014). Wearable technologies tracking health indicators, such as heart rate, are increasingly used in healthcare (Masoumian Hosseini et al., 2023). Research suggests that an increase in HR can indicate physiological or emotional arousal, such as pain, stress, and in some cases joy (Kildal et al., 2021; Raz & Lahad, 2022). Regarding individuals with communication difficulties, studies have found that sensors are needed to objectively identify distress and pain in non-verbal patients with intellectual disability (Øderud et al., 2023), and that monitoring of heart rate can identify acute distress and pain in this patient group (Kildal et al., 2021). In addition, caregivers also perceive HR monitoring as a potentially valuable tool for gaining insight and a better understanding of individuals with intellectual disability who are unable to communicate verbally (Boysen et al., 2024). As previously stated, the caregiver-patient relationship and interactions may benefit the quality of life and outcomes for the patient (Sharkiya, 2023; Hostyn et al., 2011; Petry & Maes, 2007; McGilton et al., 2012), which makes it important to know how emotional dynamics can be affected by tools like HR monitoring.

While HR monitoring may improve caregivers’ ability to detect distress, little is known about how it affects the *relational dimension of care* - specifically, caregivers’ own sense of closeness and safety in their interactions. Understanding this potential relational benefit is essential, as relational attunement and emotional connection are core components of high-quality care in PIMD. To capture both in-depth insight into the experiences of caregivers and measurable change, a mixed-methods design is chosen with three quantitative hypotheses and one qualitative aim.

## Methods

The overall objective of this study is to investigate how the use of HR monitoring as a communicative tool in everyday care affects caregivers’ perceived closeness and safety in their interactions with patients with severe communication difficulties (Fig. 1). The main research question is:

**Figure 1:**
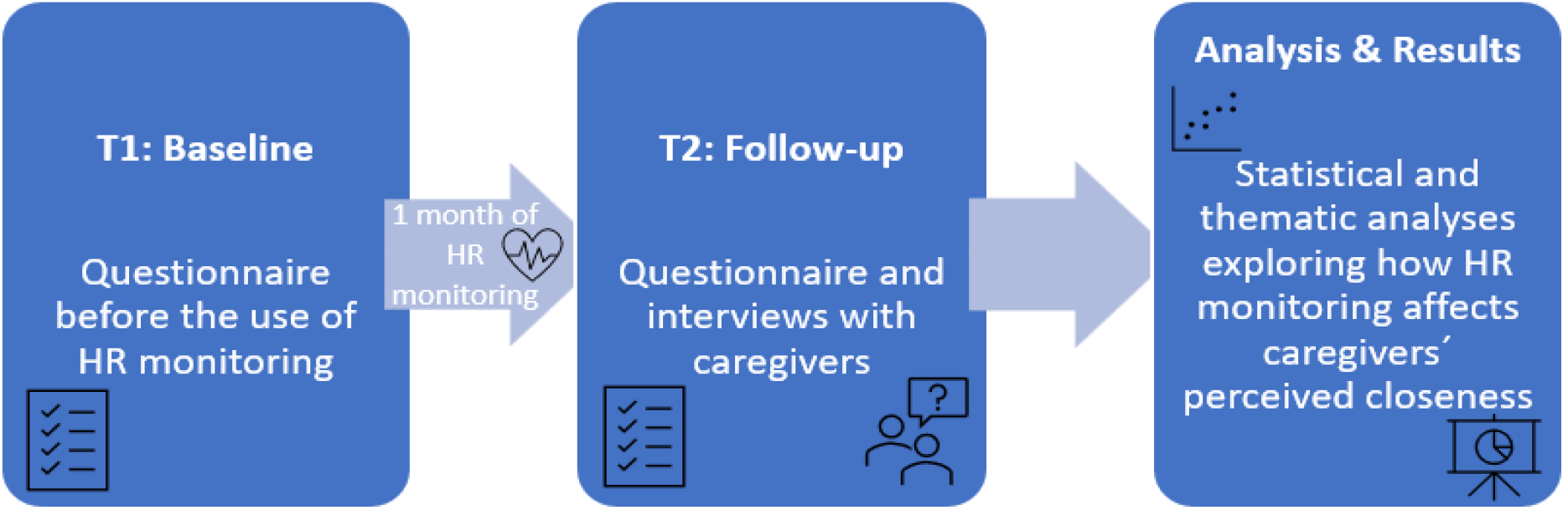
Overview of design. HR: Heart rate

How does the use of HR monitoring in everyday care affect *caregivers’ perceived closeness and safety* in interactions with patients with severe communication difficulties, *as assessed through pre-post self-report and qualitative interviews?*

### Primary Hypothesis (Quantitative)

**H1:** Caregivers will report significantly higher levels of perceived closeness in interactions with patients with communication difficulties after a month of using HR monitoring, compared to baseline (pre-HR use).

### Secondary Hypotheses (Quantitative)

**H2:** Caregivers will report changes in levels of perceived safety in interactions with patients with communication difficulties after 1 month of using HR monitoring, compared to baseline (pre-HR use).

**H3:** The increase in perceived closeness and safety from T1 (pre-HR use) to T2 (1 month of HR use) will be greater among caregivers who report more initial difficulty understanding the patient, measured via baseline questionnaire items.

### Qualitative Aim

To explore how caregivers describe the impact of HR monitoring on their sense of closeness and safety in relationships with patients with severe communication difficulties, as expressed in individual interviews.

Participants will be recruited as part of the ongoing clinical trial on HR monitoring for the detection of acute pain in non-verbal patients (Kildal et al., 2023b) registered at ClinicalTrials.gov (NCT05738278). Recruitment will take place through care institutions affiliated with the Department of Neurohabilitation at Oslo University Hospital, Norway.

The study population consists of professional caregivers employed in residential care settings for individuals with major communication difficulties. Their patients have neurodevelopmental conditions, such as ID, genetic syndromes, CP, and co-occurring epilepsy. Many have additional somatic diagnoses.

All patients fall within the category of PIMD. Communication impairments in this group include both verbal and nonverbal expression, and the severity of these impairments often prevent patients from communicating distress or pain, increasing the risk that such experiences go undetected.

### Inclusion criteria

Caregivers who work regularly with patients with communication difficulties and who participates in the clinical testing of HR monitoring in care.

### Exclusion criteria

Caregivers who do not have regular, direct interaction with patients with communication difficulties. Caregivers not involved in the HR monitoring trial.

To estimate the minimum detectable effect size for the primary hypothesis, a power analysis for a paired samples t-test was conducted using Jamovi version 2.6.44 (The Jamovi project, 2025). In the analysis, a one-tailed test was chosen, as the expected effect is directional, since it is expected that HR monitoring will increase the feeling of closeness. The minimum desired power was set to 0.8, with an alpha (α) of 0.05. The project has the capacity to recruit 58 participants. Allowing for an anticipated 15% dropout, this sample size provides sufficient power to detect effect sizes as small as *d* = 0.36 (see Figure 2). This threshold is consistent with prior studies of interventions for caregivers, which typically report small to moderate effect sizes (Terracciano et al., 2020; Heckel et al., 2018; Gitlin et al., 2010).

**Figure 2:**
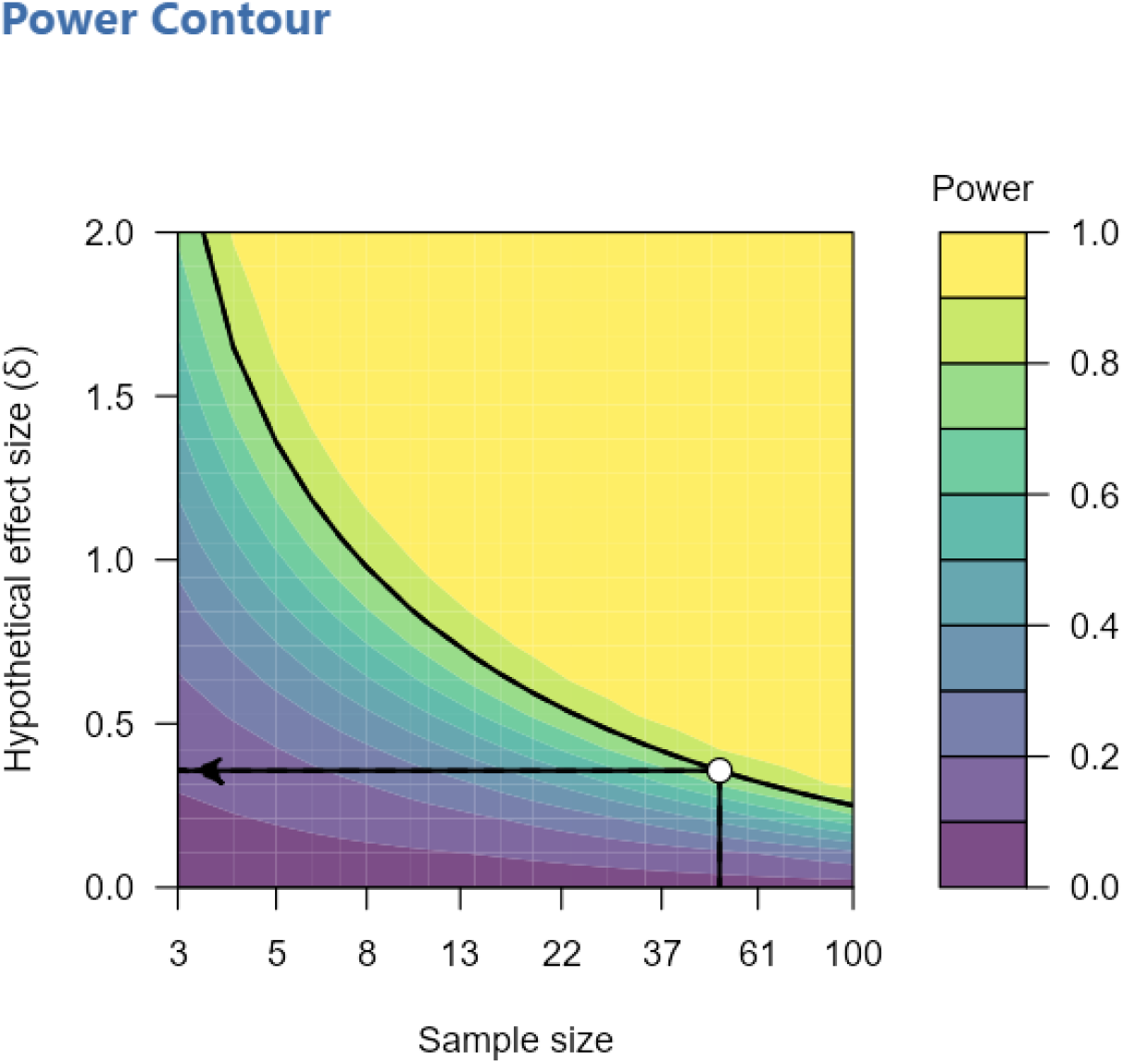
A power contour plot visualization the range of effects that can reliably detected for a range of sample sizes and hypothetical effect sizes. With an α = 0.05 (one-tailed) and a sample size of 50 participants, the study is sufficiently powered (80%) to detect effect sizes as small as 0.357

### Quantitative data collection

Caregivers will complete a questionnaire (see supplementary materials) at baseline (T1) and after 1 month of using HR monitoring in daily care (T2). The questionnaire includes: 1) Study-specific items on closeness and safety (5-point Likert scales); 2) The Non-Verbal Patient-Caregiver Understanding Questionnaire (NPUQ) assessing caregiver-perceived difficulty in understanding the patient′s communication (Kildal et al., 2023a).

The purpose of the questionnaire is to explore how caregivers report that HR monitoring affects their experience of closeness and safety in relation to the patient. The participants will answer on a Likert scale from 1-5 on the statements about closeness and sense of safety. In addition, NPUQ (Kildal et al., 2023a) will be used to assess the patient’s degree of communication difficulties (questions 6-10), and the caregivers′ experienced difficulty understanding the patient (questions 1-5).

### Measurement of perceived closeness

Caregivers will complete a brief, behaviorally anchored questionnaire designed to assess their perceived emotional closeness in one-to-one interactions with a designated patient. The items will be phrased in simple, relational terms (e.g. *“When you interact with this patient, do you feel emotionally connected?”*). Responses will be given on a 5-point behaviorally anchored Likert scale:

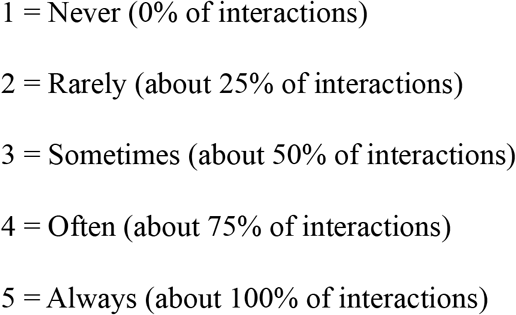

Caregivers will complete the same questionnaire at baseline (pre-intervention) and after one month of HR monitoring use (post-intervention). This allows for direct comparison of pre- and post-intervention ratings. To capture caregivers′ subjective evaluation of change and to mitigate potential ceiling effects, an additional single-item question will be included post-intervention:

> *“Compared with before using HR monitoring, my perceived closeness to this patient is: Increased / No change / Decreased”*.

### Qualitative data collection

For the qualitative data collection, interviews will be conducted with a subset of caregivers (4-6) following an interview guide with semi-structured open-ended questions (see supplementary materials). The purpose of the interviews is to explore how caregivers describe how HR monitoring affects their experience of closeness and safety in caregiving situations with the patients. The interviews will be conducted after the caregivers have used HR monitoring with patients. The interviews will last for approximately 1 hour each. The qualitative part of the study may evolve and be adjusted throughout the research process based on practical considerations and emerging insights. A reflexivity diary will also be used throughout the research process to note reasons behind choices made along the way, thoughts, decisions, reflections and potential influences of the researcher’s perspective.

### Quantitative data analysis

To analyze the quantitative data from the questionnaire, descriptive statistics with average and standard deviation (SD) for all scales at each measurement time, distribution of responses, and demographic variables will be included. It is hypothesized to be a positive correlation between the length of the caregiver-patient relationship (in months) and overall levels of reported closeness and safety, but this is not included as a hypothesis. Pearson′s r or Spearman′s rho, depending on whether the data is normally distributed or not, will be used to explore the possible correlation between the length of the caregiver-patient relationship and perceived closeness and safety. A regression analysis will be used to examine whether baseline difficulty in understanding the patient predicts change in safety and closeness. To examine potential changes in perceived safety and closeness from pre- to post-measurements, a within-subjects t-test (paired samples t-test) will be used. A one-tailed test will be used for closeness, as an increase is expected, while a two-tailed test will be used for safety, as both decreases and increases are possible. In addition, a spaghetti plot will be created for visualisation of individual trajectories.

### Qualitative data analysis

To analyze the qualitative data, thematic analysis based on Braun and Clarke will be used on the interview transcriptions (Braun et al., 2024).

### Ethical considerations

Informed consent will be obtained from all the caregivers involved in this study by providing an information letter and having them sign a consent form to ensure that they understand the project, its purpose, and that their participation is voluntary. The participants will be informed about how the data is used and stored, and their right to withdraw from the study at any time. All data will be pseudonymized to ensure anonymity. TSD will be used to store quantitative and qualitative data securely. The gender perspective is not central to the research question, but demographic variables are included to identify potential gender patterns in the data. The project is approved by The Regional Committees for Medical and Health Research Ethics South East Norway (REK), concession #2016/1956, #2009/932, and #2012/1967. The study is conducted in accordance with the Declaration of Helsinki (World Medical Association, 2001).

## Discussion

The study may contribute to the understanding of how HR monitoring affects relational aspects of care. Previous research has shown that HR monitoring can be used to gain a better understanding of individuals with intellectual disability (Boysen et al., 2024), and to identify pain and distress in nonverbal patients (Øderud et al., 2023; Kildal et al., 2021), but currently little is known about how HR monitoring affects the caregivers experiences and perceptions of closeness and safety in the interactions and relationships with patients with PIMD. The study moves beyond including only the physiological aspects of HR monitoring, by exploring it as a potential influence on relational quality and emotional attunement.

By combining both qualitative and quantitative methods, we might get more insight and a deeper understanding of how HR monitoring affects relational aspects of care. The quantitative questionnaire will result in measurable data on closeness and safety, while the qualitative interviews might add possible nuances and contexts to the caregivers’ experiences. Together, these methods might highlight both potential challenges and benefits of using HR monitoring in care.

The study might also result in practical implications for future technological tools and interventions made to improve the quality of life for patients with PIMD, prevent uncertainty and support caregivers. In addition, the study may result in more knowledge in the context of PIMD, that may be used to inform theoretical perspectives on emotional connection and relational care.

At the same time, the study has several possible limitations worth considering. First, we might not get as many participants as we want, which may lead to a small sample size. A smaller sample size might affect statistical power and the generalizability of the results. It should be noted that the findings may not be generalizable to other care settings or caregivers.

Second, the results are based on self-reported, subjective experiences both in the questionnaire and in the interviews. We do not have any observational or objective validation of the measurements of the feelings of safety and closeness. This might heighten the risk of recall bias, individual variations of interpretations of closeness and safety, or the participants answering what they believe is socially desirable, or answering what they think the researcher would want them to answer.

Third, there might be a possible bias in what kind of caregivers choose to answer both in the interviews and the questionnaire. For instance, they might be more positively oriented towards technology, score higher on openness to new experiences or have different motivations or attitudes compared to those who do not wish to participate, which can affect the results of the study.

Fourth, contextual factors are not controlled for. Contextual factors, for instance a caregiver’s workload or the culture within the workplace can affect and shape a caregiver’s experience of closeness and safety. Finally, the study cannot determine causality between HR monitoring and relationship outcomes, but it has an explorative value that might raise important questions or indicate tendencies. The study may provide a possible basis for future and larger-scale research.

## Conclusion

This is a mixed methods study exploring how HR measurements affect caregivers’ sense of closeness and safety with patients with severe communication difficulties. It is anticipated that this research will increase knowledge of how closeness and safety is affected by HR measurements.

## Abbreviations

PIMD: Profound intellectual and multiple disabilities;
ID: intellectual disability;
CP: cerebral palsy;
HR: heart rate;
NPUQ: The Non-Verbal Patient-Caregiver Understanding Questionnaire;
The clinical trial: the clinical trial of heart rate monitoring to detect acute pain in non-verbal patients (registered at ClinicalTrials.gov NCT05738278).

## Data availability

Not applicable. As this is a preregistration protocol, no data have been collected or analyzed yet.

## Declaration of conflicting interest

No conflicts of interest. The authors and their institutions have not received any payments or services from a third party in the last 36 months that could give the appearance of potentially influencing or be perceived as influencing the submitted work.

## Funding

Not applicable. The authors and their institutions have not received any payments or services from a third party at any time for any aspect of the submitted work.

## Supplementary materials

Supplementary materials can be found on the project’s Open Science Framework page: https://osf.io/t6g7d/

## Notes

### Competing Interest Statement

The authors have declared no competing interest.

### Funding Statement

This study did not receive any funding

### Author Declarations

The project is approved by The Regional Committees for Medical and Health Research Ethics South East Norway (REK), concession #2016/1956.

